# “You can change your life through sports”: Physical activity interventions to improve the health and wellbeing of adults experiencing homelessness: a mixed methods systematic review

**DOI:** 10.1101/2023.07.26.23293203

**Authors:** Jo Dawes, Raphael Rogans-Watson, Julie Broderick

## Abstract

**Objectives:** Systematically synthesise evidence of physical activity interventions for people experiencing homelessness (PEH).

**Design:** Mixed-methods systematic review.

**Data sources:** EMBASE, Web of Science, CINAHL, PubMed (MEDLINE), PsycINFO, SPORTDiscus, and Cochrane Library, searched from inception to October 2022.

**Eligibility Criteria:** PICO framework: Population (quantitative and qualitative studies of PEH from high-income countries); Intervention (physical activity, any setting); Comparison (with/ without comparator); Outcome (any health/wellbeing-related outcome).

**Results:** 3,614 records screened, generating 17 reports [16 studies, 11 qualitative and 5 quantitative (1 RCT, 3 quasi-experimental, 1 analytical cross-sectional)] from UK, USA, Denmark and Australia, including 539 participants (501 PEH, 38 staff). Interventions: soccer (n= 6), group exercise [indoor (n=3), outdoor (n=5)] and individual activities (n=2). Risk of bias assessed using JBI critical appraisal tools. Mixed methods synthesis identified physical and mental health benefits. Qualitative evidence highlighted benefits carried into wider life, challenges participating and positive impact on addiction. Qualitative and quantitative evidence was aligned for the mental health benefits of outdoor exercise and increase in physical activity from indoor group exercise. Quantitative evidence suggested improved bone health and blood lipid markers.

**Conclusion:** Diverse interventions were identified with soccer predominating. Qualitative evidence suggested physical activity can benefit health and wellbeing with positive translation to wider daily life. There was some positive quantitative evidence, although most was inconclusive. Evidence suggests a tentative recommendation for physical activity interventions for PEH, however a limitation is that results may not be transferable outside high-income countries. More high-quality research is required to determine effectiveness and optimal programme design.

**What is already known?:** People experiencing homelessness suffer a disproportionally higher burden of physical and mental health conditions than housed populations.

Regular physical activity can address many health conditions prevalent amongst people experiencing homelessness.

**What are the new findings?:** There is evidence of a variety of physical activity interventions that have been designed and provided to engage people experiencing homelessness (for example: soccer, outdoor and indoor group activities, and individual activities).

The synthesis of qualitative and quantitative evidence suggests that physical activity can benefit the mental and physical health of people experiencing homelessness with positive translation of benefits to wider life.

## INTRODUCTION

Homelessness is an extreme form of social exclusion[1, 2] related to poverty in developed countries.[3] People experiencing homelessness (PEH) are defined as those who are ‘roofless’ (e.g. no fixed abode) and ‘houseless’ (e.g. living in hostel, shelter, temporary accommodation) in accordance with the European Federation of National Organisations Working with the Homeless (FEANTSA).[4] Prior to the COVID-19 pandemic, homelessness in the United Kingdom (UK) had increased annually since 2010[5] with estimates of all categories of homelessness in England standing at 280,000 people,[6] of which 4,266 were estimated to be sleeping on the streets.[7]. The Organisation for Economic Co-operation and Development (OECD) estimates almost 2 million people are experiencing homelessness in 35 OECD countries.[8].

PEH have poorer health than the general population,[9, 10] often characterised by a tri-morbidity of mental health diagnoses, chronic physical health conditions and addiction.[9] Poor health is thought to be both precipitated and exacerbated by poor living conditions, lack of resources, social exclusion, stigmatisation and difficulty accessing suitable health services.[11]

Physical activity is beneficial for people with disabilities and chronic health conditions. Guidance suggests the type and amount of physical activity should be determined by a person’s abilities and the severity of their condition or disability, which may change over time.[12] PEH live with a high burden of physical deficits,[13] falls and frailty,[14] respiratory disease, cardiac problems, stroke and diabetes,[15] which could be positively influenced by physical activity. Moreover, research suggests that PEH may be less physically active,[16] so may miss out on health gains and reduced risk of harm that physical activity affords people with these conditions. It is important that this population have opportunities for physical activity to stabilise or reverse physical declines associated with homelessness. Given the multiple barriers PEH face accessing services, it may be important that physical activity interventions are specifically tailored to their needs to optimise reach and participation. This research poses two research questions: what are the range of physical activity interventions provided to PEH? And, what is the evidence supporting effectiveness of these interventions?

### Aims

This review aims to summarise the available evidence for physical activity interventions intended to improve health outcomes of adults experiencing homelessness, focusing on physical activity interventions and their effectiveness for improving health outcomes.

## METHODS

### Design

A preliminary scoping review revealed that published literature in the field of physical activity for PEH comprised quantitative and qualitative research, therefore a mixed methods systematic review was adopted. This allow for the findings of effectiveness (quantitative evidence) and participant experiences (qualitative evidence) to be brought together, to facilitate broader understanding of whether and how interventions worked.[17, 18] This systematic review was conducted following the Preferred Reporting Items for Systematic Reviews and Meta-analyses (PRISMA) 2020 guidelines[19] and checklist[20] and the protocol was registered a priori in PROSPERO database (reference number: CRD42020216716).

### Identification

#### Defining search terms

Initial search terms were generated by reviewers (JD, RRW and JB), who between them, have extensive clinical and research expertise and experience in the health of PEH, physical activity and systematic review methodology. Search terms were refined and tailored for a preliminary search of Medline, and used to test proof of concept and search strategy. The search syntax (Supplementary File 1) was designed by a professional librarian, in collaboration with two reviewers (JD and RRW).

### Searches

Search terms were refined, adapted and run in Medline, EMBASE, Web of Science, CINAHL, PsycINFO, SPORTDiscus, and the Cochrane Library. Searches were conducted on 17^th^ Feb 2021, including literature from the previous 30 years (1991-2021) restricted to English language only. The searches were re-run using original search terms, by a specialist librarian at Trinity College Dublin on 19^th^ Oct 2022 to identify any new reports published between dates 21^st^ Oct 2021 to 19^th^ Oct 2022. All previous databases were searched, except SPORTDiscus, as it was not unavailability in the institution’s library databases. Duplicates were removed at this stage. Reference lists of relevant systematic reviews were hand searched for reports to be added for screening. Additionally, reference lists of all included studies were hand searched for relevant studies. Corresponding authors of records that comprised an abstract only were contacted where possible to request full text reports.

### Screening

#### Title and abstract screening

On completion of the identification process, all report titles and abstracts were uploaded to the online systematic reviewing management system Covidence. Two pairs of reviewers (JD/RRW and JD/JB) independently performed (i) title and abstract screening and (ii) full text screening, judged against predetermined protocol criteria. In the event of disagreement, the third reviewer (JB or RRW) was consulted for an additional opinion.

The PICO framework was used to identify inclusion criteria. For inclusion, all following criteria were to be met:

#### Population

Studies that included adults who were homeless under the ETHOS criteria for homelessness[4], that is rooflessness, houselessness, living in insecure housing, or living in inadequate housing. Age >18 years.

#### Intervention

Studies that included any physical activity intervention delivered as a stand-alone intervention or part of multimodal intervention, in any setting. Studies undertaken in high income countries[21] were included, where there is assumed consistency in health and social care infrastructure as well as in family and community support systems, which impact on how homelessness is perceived and managed.[22]

#### Outcome

This mixed methods review included quantitative studies reporting any measures demonstrating health outcomes, including but not limited to, primary measures such as cardiovascular fitness and strength and qualitative findings describing participant perceptions linking physical activity intervention to health and/ or wellbeing outcomes.

#### Comparison

The presence of a comparison group was not required as an inclusion criterion.

#### Study types

This review considered quantitative, qualitative and mixed methods studies.

#### Risk of bias assessment

In recognition of the diverse study designs included in this review, the JBI critical appraisal tool portfolio was a key resource for judging quality and risk of bias.[23, 24] These tools provide a criterion based checklist for determining presence (yes), absence (no), a lack of clarity (unclear) or a lack of applicability (not applicable) of quality in studies across a variety of methods.[25] To determine dependability and credibility of qualitative reports, their ConQual ratings were calculated.[26] Although Munn et al discourage cut-off values in determining quality level in quantitative studies, for clarity and consistency of this mixed methods review a pragmatic decision was made to select cut off of <25% (very low), <50% (low), <75% (moderate) and >75% (high). Munn et al state that if cut offs are preferred, these thresholds are best decided by the reviewers themselves.[25] Summary of quality assessment of all reports are summarised in Supplementary File 2.

#### Data extraction

The following data was extracted to an excel spreadsheet: study design, inclusion criteria, participants (description, number, accommodation, age, education, employment, ethnicity, race, biological sex, mental health and physical health), intervention (setting, frequency, intensity, time, type, group or individual, presence of other non-physical activity intervention components), quantitative outcome measures and qualitative themes.

Initially, JD carried out and collated data extraction of five reports. This was reviewed by RRW and JB to ensure accuracy and consistency. Once all three team members agreed the data extraction process, the remaining reports were divided between the team for completion of data extraction. Data from each report was checked for accuracy by one other member of the research team. Any inconsistencies of interpretation or reporting were discussed, and consensus reached.

#### Strategy for mixed methods data synthesis

The synthesis followed the JBI methodology for mixed methods systematic reviews,[27] whereby established convergent, segregated, results-based mixed methods frameworks for systematic reviewing were employed. [28, 29] Firstly, qualitative and quantitative data were meaningfully categorised by JD and JB respectively. Each reviewer conducted their analysis separately, independently and concurrently. JD adopted thematic analysis to synthesise qualitative data (details of themes are outlined in supplement 3). Due to the heterogeneity of quantitative studies, it was not possible for JB to carry out meta-analysis, so narrative synthesis was used. Quantitative findings were then “qualitized” to transform them into a qualitative, descriptive format. Next, quantitative and qualitative evidence were linked and organised to produce an overall ‘configured analysis’[27] and reported as a series of tables and combined narrative synthesis.

#### Equality and diversity statement

Our author and librarian team consisted of three women and two men. The author team included early and mid-career researchers and clinicians across two disciplines (medicine and physiotherapy) from two countries (UK and Ireland). This research explores physical activity interventions for people experiencing homelessness, an under-served, often marginalised and excluded population who experience extreme socioeconomic disadvantage. This population are known to have complex and chronic health needs and are an often-overlooked group in physical activity research.

## RESULTS

### Study selection

13,736 records were identified through searches. After removal of duplicates (n= 10,122) 3,614 records were screened by title and abstract, with 3,496 records excluded at this stage. 118 reports were sought for full text review, four could not be found, so 114 full text reports were reviewed-97 records were excluded at this stage (exclusions based on: 1 duplicate, 9 population, 59 intervention, 8 non-English language, 19 insufficient data, 1 protocol only). Finally, 17 reports were included for quality checking. Two reports described different aspects of a single study. Therefore, data was extracted from 17 reports describing 16 studies. The full identification, screening and inclusion process are outlined in a PRISMA diagram (Figure 1).

**Figure 1:**
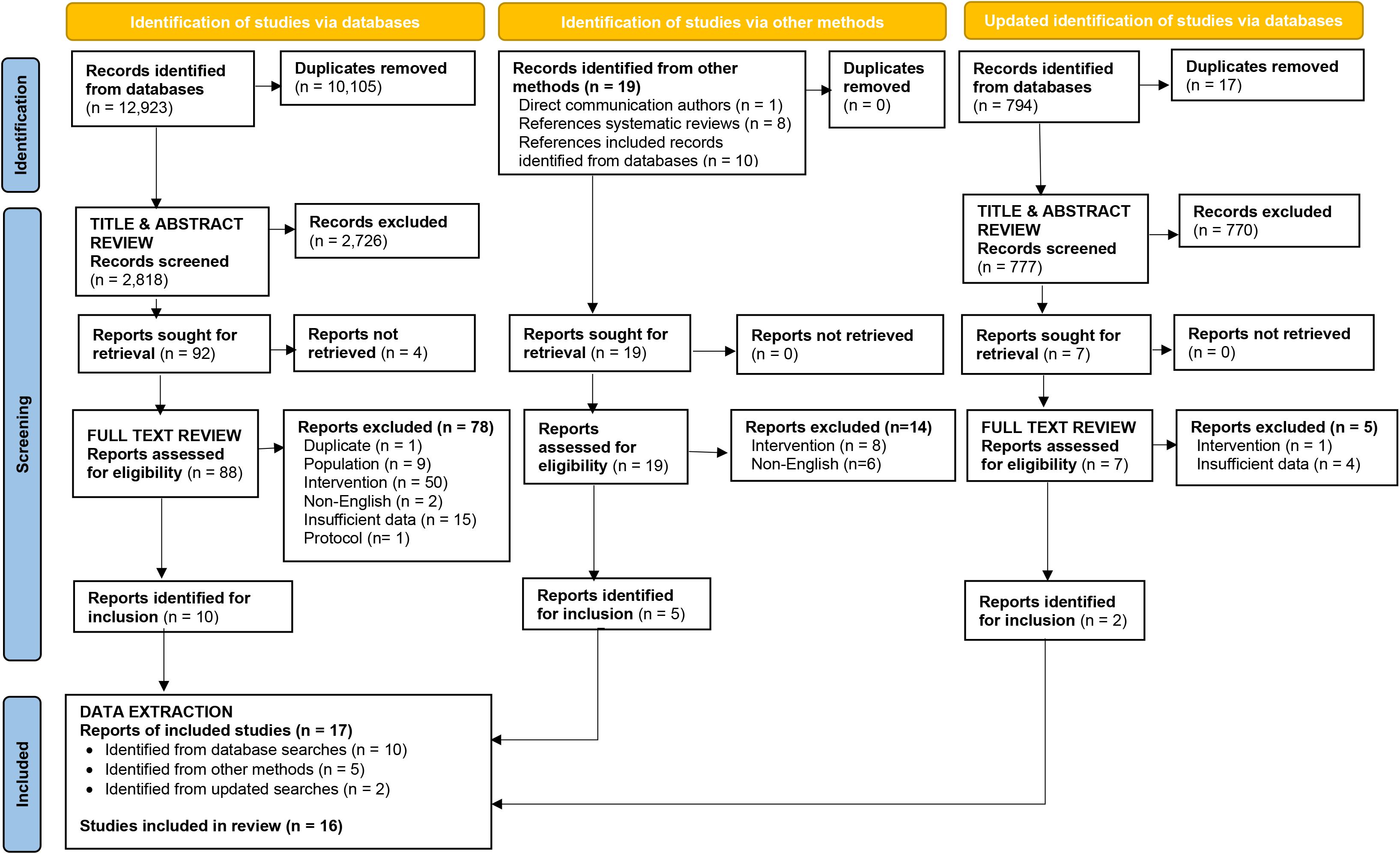
PRISMA flow diagram

### Quality assessment

The majority of the eleven qualitative studies were high quality, with eight reporting at least seven out of ten quality criteria on the JBI checklist for qualitative studies (Supplementary File 2). One study was of very low quality, [30] with only the statement of researcher positionality being clear, and all other criteria either unreported or unclear. Of the quantitative studies, the one randomised controlled trial (RCT)[31] was assessed as moderate quality due to methodological limitations, e.g. lack of clarity regarding blinding of assessor and whether treatment groups were concealed. The analytical cross-sectional study was of moderate quality, and in general, quasi-experimental studies were of high quality.

### Description of studies

Seventeen reports, describing 16 studies were included (Table 1). Of these studies, seven were from the USA, five from the UK, two from Denmark and two from Australia. The variety of designs across these studies comprised eleven qualitative and five quantitative reports (three quasi-experimental, one RCT, and one analytical cross-sectional). The interventions addressed varied, including: soccer (n=6); group outdoor exercise (n=5); group indoor multimodal exercise (n=3); and individual multimodal interventions (n=2) (Supplementary file 4).

**Table 1:** Summary of included studies.

| Study | Location | Study design | Description of homelessness/<br>accommodation | Inclusion criteria/Description of Participants | Number of<br>Participants | Intervention |
| --- | --- | --- | --- | --- | --- | --- |
| Dawes et al, 2019 | UK | **Qualitative** (interview) | No fixed abode, living in hostel, temporary accommodation, and permanent accommodation after being homeless | Attendees of one of two park-based running groups operated by the charity 'A Mile in her Shoes' in London for women defined as homeless | 11 | **Outdoor Running** (groups) |
| Grabbe et al, 2013 | USA | **Qualitative** (interview) | No fixed abode or living in shelters | Participation in at least eight gardening sessions in a daytime shelter for homeless women in a large south-eastern U.S. city | 8 | **Outdoor Gardening** (groups) |
| Grimes & Smirnova, 2020 | USA | **Qualitative** (interview) | Temporarily housed and most had experienced two or three episodes of homelessness | Men experiencing homelessness who has completed the earn-a-bike programme in the previous year in Kansas | 16 | **Mixed individual intervention** (bicycle provision, cycle safety and maintenance training) |
| Helge et al, 2014 & Randers et al, 2012 | Denmark | **Quasi-experimental** (Non-randomised controlled intervention study) | Recruited from homeless shelters and unemployment offices. | Men experiencing homelessness who were accessing services in shelters and unemployment offices in Copenhagen | 28 | **Soccer** (group training) |
| Kendzor et al, 2017 | USA | **Randomised Controlled Trial** | Living in a transitional shelter for people experiencing homelessness | 1) at least 18 years of age, 2) willing and able to attend study visits, 3) score $\geq 4$ on the Rapid Estimate of Adult Literacy in Medicine—Short Form (REALM-SF) indicating > sixth-grade literacy level, 4) physically ambulatory, 5) residents of a transitional shelter in Dallas, Texas (able to show an ID badge), and 6) had been living in the transitional shelter for $\leq 3$ months | 32 | **Mixed individual intervention (pedometer with step count goals, health education, provision of fruit and vegetables)** |
| Knestaut et al, 2010 | USA | **Qualitative** (self-reported form, debrief and journal) | People living in homeless shelter | Adults who had taken part in shelter-based dance programme | 11 | **Indoor multimodal exercise** (Instructor-led dance group) |
| Magee & Jeanes, 2013 | UK | **Qualitative** (interview) | Living in social services accommodation or hostels, and spent time with no fixed abode | Male participants of the UK squad who attended the inaugural Homeless World Cup | 6 | **Soccer** (group training and Homeless World Cup participation) |
| Malden et al, 2019 | UK | **Qualitative** (interview) | Living in hostel accommodation or on the streets | Participants of the Street Fit Scotland intervention who attended both the fitness classes and peer support components of the intervention in March 2016 | 10 | **Indoor multimodal exercise** (Instructor-led, leisure centre-based group, with peer support) |
| Norton et al, 2020 | USA | **Quasi-experimental** (non-equivalent groups longitudinal design) | Living at a shelter for women without housing | Women residents of a shelter in a large city in Texas, U.S. who had participated in the HOPE Adventure Therapy pilot program. | 82 | **Outdoor Adventure** (groups) |
| Parry et al, 2021a | UK | **Qualitative** (realist evaluation: interviews, focus groups and diary room) | Young people experiencing or at risk of homelessness (no further details provided) | Young people with experience or at risk of homelessness who attended at least one MST4Life programme workshop (Phase 1) during the time of data collection and housing staff with one-to-one experience of working with the MST4Life participants | 30 PEH<br>6 housing service staff | **Outdoor Adventure** (groups) |
|  |  |  |  |  | 5 OAE staff |  |
| Parry et al, 2021b | UK | **Qualitative** (diary room) | Living in supported accommodation for young people experiencing homelessness | Young people with experience or at risk of homelessness who attended and engaged in Phase 1 of MST4Life from different cohorts of MST4Life over a 5-year period (2014-2019) | 54 | **Outdoor Adventure** (groups) |
| Randers et al, 2018 | Denmark | **Analytical cross-sectional** (intervention study) | Described as homeless, no further details provided | Women experiencing homelessness from three countries (Denmark, Norway and Belgium) who participated in 4-a-side street soccer at Women's Homeless World Cup in Amsterdam in December 2015 | 15 | **Soccer** (Homeless World Cup tournament participation) |
| Sherry, 2010 | Australia | **Qualitative** (interview) | Experience of homelessness in the preceding two years or were participating in a drug or alcohol rehabilitation programme | Team members of the 'Street Soccerroos', the Australian Homeless World Cup team and | 8 | **Soccer** (group training and Homeless World Cup participation) |
| Sherry & Strybosch, 2012 | Australia | **Qualitative** (Ethnographic case study) | Past or current experience of homelessness and associated social disadvantage | Team members of Australia's Community Street Soccer Programme (CSSP) over a four-year period, which is for people who are homeless or experience of homelessness/social disadvantage, staff from the CSSP and key stakeholder or support workers involved in the development and delivery of the programme | 165 players<br>11 Coaches<br>10 (support workers) | **Soccer** (group training) |
| Shors et al, 2014 | USA | **Quasi-experimental** (intervention study) | "Recently homeless" with experience of poverty, trauma, or addictive behaviours | Young mothers who were recently homeless then "rescued from the streets" and given housing and food in a residential center, where they lived with their children | 15 | **Indoor multimodal exercise** (Instructor-led meditation and choreographed aerobic group exercise) |
| Welty Peachey et al, 2013 | USA | **Qualitative** (focus groups) | Recruited from a tournament for homeless individuals. So, assumption that all participants were homeless – no further details supplied | Players and coaches who 'represented the geographic areas' of the U.S.-based Street Soccer USA Cup | 11 players<br>6 staff | **Soccer** (National Cup tournament participation) |

### Study populations

Supplementary File 5 provides detail of each study population included in this systematic review. Across the 16 studies, 501 PEH were participants. Some studies included women only (n=5), men only (n=4) or mixed cohorts (n=7). Three qualitative studies reported staff/ coaches’ perspectives (n=38). The age range of participants who were homeless was 16 – 65 years. It was specified in the review protocol that only studies with participants >18 years would be included. However, for pragmatic reasons, several studies[32–36] were included despite containing participants from the age of 16 years. In these studies, proportions of participants <18 years was not specified, although one study[35] stated the ‘majority’ of participants were between the ages of 20-24 years. Descriptions of study participants’ experiences of homelessness varied but were mainly focused on: street homeless, living in hostel/ shelter, transitional/ social service accommodation, or “homeless at time of intervention”. Studies that focused on Street Soccer and the Homeless World Cup invited participation from PEH and other socially excluded groups, for example, people attending unemployment offices or drug rehabilitation services. Although these studies did not define proportions of participants experiencing homelessness, for pragmatic reasons, they were included, as the intervention had been specifically designed for PEH.

### Physical activity interventions and their components

Supplementary File 4 provides a description of all included interventions. Studies included: six soccer interventions [tournament focused (n=2), group training focused (n=2), and combining group training and tournament participation (n=2)]; five group outdoor exercise [adventure training (n=3), running (n=1) and gardening (n=1)]; three group indoor multimodal exercise (aerobic-based circuits (n=2) and dance (n=1)]; and, two individual multimodal interventions (pedometer with step goals and earn-a-bike scheme). Supplementary file 4 also provides programming variables, including: setting; frequency; intensity; time; type; and, the presence of other non-physical activity components of multimodal interventions.

### Soccer

Six studies investigated the impact of soccer for PEH. These studies (three reports) explored soccer group training (n=2),[35, 37, 38] tournament participation (n=2) [36, 39] and interventions of training for and participating in tournaments (n=2.)[32, 40] The studies involving tournaments were focussed around national or international tournaments such as the Homeless World Cup or Street Soccer USA Cup.[32, 36, 39, 40]

### Group outdoor exercise

Five studies provided evidence of the value of group outdoor exercise. These included group outdoor adventure (n=3),[33, 34, 41] women’s running groups (n=1),[42] and women’s gardening groups.[43] These studies described multi-modal interventions, including outdoor adventure interventions which contained multiple activities (e.g. archery, rock climbing, hiking) and all studies reported additional support, such as the provision of education, debriefing, opportunities for reflection, childcare, food or clothing.

### Group indoor multimodal exercise

All group indoor multimodal exercise studies (n=3) were instructor-led interventions provided to small groups in settings such as leisure centres,[44] or shelter recreation rooms.[30] All studies were multimodal as they combined different types of activity, e.g. stretching, cardiovascular exercise, varied dance genres, aerobic circuits, strength-based exercise to music and meditation.

### Individual multimodal interventions

Two studies reported interventions for individuals.[31, 45] One involved participants wearing a pedometer and working towards a step goal-this was provided along with an educational newsletter and fruit and vegetable snacks.[31] The other study described cycle training to learn road safety and cycle maintenance, alongside earning a bicycle for individual use.[45]

### Intervention and Outcomes

Findings are described across four tables (Tables 2-5). Table 2 shows all synthesised findings relating to mental health and Table 3 shows all synthesised findings relating to physical health where the configured analysis identified qualitative and quantitative evidence supporting matched themes. Table 4 shows evidence that was identified in either quantitative or qualitative reports alone. For example findings where only quantitative data existed was related to bone health and blood markers. Whereas qualitative evidence only was identified relating to other important aspects of physical activity, not specifically or directly health related, such as the benefits carried into wider life, challenges of participation and addiction.

**Table 2:** Summary of synthesised findings relating to mental health benefits of physical activity participation.

| Qualitative |  |  | Quantitative |  |  | Synthesis |
| --- | --- | --- | --- | --- | --- | --- |
| Theme | Sub theme | Level of confidence in evidence for intervention type | Description of outcome and measurement tool | Outcome data | Level of confidence in evidence for intervention type | Overarching synthesised finding |
| Belief that physical activity facilitated people experiencing homelessness in self-development and their ability to cope with life situations | Confidence, empowerment and self-esteem | **High** (Group running[42], Soccer-group training and tournament participation[32], Indoor group instructor-led exercise[44])<br>**Moderate** (Earn-a-bike[45], Group outdoor adventure[33])<br>**Low** (Gardening group[43], Soccer-group training[35])<br>**Very low** (Group instructor-led dance[30]) | No change to goal directed energy, measured using The Hope scale (agency subscale) | No significant difference between intervention and control groups | **Moderate** (Group outdoor adventure[41]) | High quality qualitative evidence that group running, soccer and indoor group exercise and moderate quality qualitative evidence that group outdoor adventure and Earn-a-bike enhance confidence, empowerment, and self-esteem. However, quantitative evidence does not support this. |
|  | Resilience, coping and hope | **High** (Group running[42], Group outdoor adventure[34])<br>**Moderate** (Group outdoor adventure[33])<br>**Low** (Gardening group[43], Soccer-group training[35])<br>**Very low** (Group instructor-led dance[30]) | No change to planning and accomplishing goals, measured using The Hope scale (pathway) | No significant difference between intervention and control groups | **Moderate** (Group outdoor adventure[41]) | High quality qualitative evidence that group running and group outdoor adventure enhances resilience, coping and hope, but quantitative evidence does not support this. |
| | Independence, focus, personal development and relationships | **High** (Group running[42], Soccer-group training and tournament participation[32])<br>**Moderate** (Group outdoor adventure[33], Earn-a-bike[45])<br>**Low** (Gardening group[43], Soccer-group training and tournament participation [40]) | Positive changes in life functioning, measured by (Outcomes rating scale), including interpersonal and social wellbeing | Significant difference between IG and CG for social wellness ($p < 0.01$) | **Moderate** (Group outdoor adventure[41]) | High quality qualitative evidence that group running, and soccer and moderate quality qualitative evidence that group outdoor adventure and Earn -a-bike enhance independence, self-regulation and personal development. This is supported by moderate quality quantitative |
|  |  | **Very low** (Group instructor-led dance[30]) |  |  |  | evidence that outdoor adventure improves life functioning. |
| Belief that physical activity resulted in people experiencing homelessness feeling mentally better | Positive effect on stress and anxiety | **High** (Group running[42], Indoor group instructor-led exercise[44], Group outdoor adventure[34])<br>**Moderate** (Soccer- tournament participation[36], Earn-a-bike[45])<br>**Low** (Gardening group[43], Soccer-group training and tournament participation [40])<br>**Very low** (Group instructor-led dance[30]) | Positive changes to flow and worry, measured by the Flow Kurtz Scala (Flow Short scale) | People who anticipated in soccer tournament scored highly for flow ($5.5 \pm 0.8$) whereas the score for worry was moderate ($4.6 \pm 1.3$) | **Moderate** (Soccer-tournament participation[39]) | High quality qualitative evidence that group running, indoor group exercise and outdoor adventure and moderate quality qualitative evidence that soccer and Earn-a-bike have a positive effect on stress and anxiety. This is not supported by quantitative evidence. |
|  |  |  | Decreased anxiety levels, measured by Beck Anxiety Inventory | A significant decrease in anxiety in intervention group, but p-value not supplied | **Low** (Indoor group aerobic-based dance[46]) |  |
| | Positive effect on mood and state of mind | **High** (Group running[42], Soccer-group training and tournament participation[32], Indoor group instructor-led exercise[44]), Group outdoor adventure[34])<br>**Moderate** (Earn-a-bike[45])<br>**Low** (Gardening group[43])<br>**Very low** (Group instructor-led dance[30]) | Positive changes in life functioning, measured by (Outcomes rating scale), including personal and overall wellbeing | An increase in overall wellbeing at a faster rate in intervention group compared to control group ($p < 0.05$) | **Moderate** (Group outdoor adventure[41]) | High quality qualitative evidence that group running, soccer and indoor group exercise and moderate quality qualitative evidence that Earn-a-bike enhances mood and state of mind. This is supported by moderate quality quantitative evidence that group outdoor adventure improves wellbeing. |
|  |  |  | Decreased depression levels, measured by Beck Depression Inventory | Significant decrease in depression in intervention group, p-value not supplied | **Low** (Indoor group aerobic-based dance [46]) |  |

**Table 3:** Summary of synthesised findings relating to physical health benefits of physical activity participation.

| Qualitative |  |  | Quantitative |  |  | Synthesis |
| --- | --- | --- | --- | --- | --- | --- |
| Theme | Sub theme | Level of confidence in evidence for intervention type | Description of outcome and measurement tool | Outcome data | Level of confidence in evidence for intervention type | Overarching synthesised finding |
| Belief that physical activity improves the body shape of people experiencing homelessness | Improved body shape and self-image | **High** (Running groups[42], Indoor group instructor-led exercise[44]) | Positive changes to body composition, measures using x-ray absorptiometry (DXA) | Weightbearing fat mass decreased by 13% from 14.0±7.2kg to 12.2±6.5kg. This change was significantly different (p=0.008) in the intervention group compared to the control group. | **High** (Soccer-group training[37]) | High quality qualitative evidence that group running and indoor group exercise can improve body shape and weight loss. This is supported by high quality quantitative evidence that soccer significantly decreases fat mass. |
|  | Weight loss | **High** (Running groups[42])<br>**Low** (Soccer- group training[35]) |  | Total fat mass decreased by 13% from 1.7 kg (95% CI: -2.6 to -0.9 kg over 12 weeks. This change was significantly different (p< 0.01) in the intervention group compared to the control group. | **High** (Soccer-group training[38]) |  |
| Belief that physical activity improves physical condition of people experiencing homelessness | Perception of improved fitness levels | **High** (Running groups[42], Soccer- group training and Tournament participation[32], Indoor group instructor-led exercise[44])<br>**Moderate** (Earn-a-bike[45])<br>**Low** (Soccer- group training[35])<br>**Very Low** (Group instructor-led dance[30]) | Positive changes to cardiovascular fitness, measured using cycle ergometer test to exhaustion | Maximal oxygen uptake was elevated from 2.69 ± 0.47 to 2.95 ± 0.52 L O<sub>2</sub> min<sup>-1</sup> (95% CI: 6.7 to 13.1%) after 12 weeks. This change was significantly different (p= 0.01) in the intervention group compared to the control group. | **High** (Soccer-group training[38]) | High quality qualitative evidence that group running, soccer and indoor group exercise and moderate quality qualitative evidence that earn-a-bike improves fitness levels. This is supported by high quality quantitative evidence that soccer improves cardiovascular fitness and endurance. |
|  |  |  | Positive changes to cardiovascular fitness, measured using yo-yo endurance test | Endurance was improved by 45% which was 81 s (1,034 ± 218 to 1,116 ± 225 s, 95% CI: 47–128) s after 12 weeks. This change was significantly different (p= 0.05) in the intervention group compared to the control group. | **High** (Soccer-group training[38]) |  |
|  |  |  | Positive changes to cardiovascular | There was a significant pre-post difference in maximal oxygen consumption (VO<sub>2</sub> peak) in the | **Low** (Indoor group aerobic-based dance) |  |
|  |  |  | fitness, measured using submaximal treadmill exercise test | intervention group (p=0.05). No between group comparison was available. |  |  |
|  | Physical skill development | **Moderate** (Group Outdoor adventure[33])<br>**Low** (Soccer- group training[35])<br>**Very Low** (Group instructor-led dance[30]) | No changes to postural balance, assessed by single legged flamingo balance test | No difference between pre-post intervention changes in postural balance between intervention and control groups | **High** (Soccer-group training[37]) | Moderate quality qualitative evidence that group outdoor adventure improves physical skill development, this was not backed up by quantitative findings amongst a soccer cohort. |
| Belief that participating in physical activity interventions makes people experiencing homelessness more active in general | Participating in the physical activity intervention increased physical activity in everyday life | **High** (Running groups[42], Indoor group instructor-led exercise[44])<br>**Moderate** (Earn-a-bike[45])<br>**Low** (Soccer-group training[35]) | Positive changes to physical activity levels, as measured by accelerometry | Minutes/ day of moderate and vigorous physical activity was greater among participants in the intervention group (median 60 minutes/day) compared to the control group (median 41 minutes/day) (p=0.00036) | **Moderate** (Pedometer with step-count[31]) | High quality qualitative evidence that group running and indoor group exercise and moderate quality qualitative evidence that Earn-a-bike can positively influence physical activity levels amongst intervention participants. This is supported by moderate quality quantitative evidence that a pedometer with step-count goal significantly increased moderate/ vigorous minutes per day of physical activity per day. |
|  | Broader aspects of the intervention (clothes, equipment, skill) facilitated physical activity participation | **High** (Running groups[42])<br>**Very Low** (Group instructor-led dance[30]) |  |  |  |  |

**Table 4:** Outcomes where quantitative only or qualitative only findings exist, no mixed methods synthesis.

| Qualitative findings |  |  |
| --- | --- | --- |
| Theme | Sub theme | Level of confidence in evidence for intervention type |
| Belief that the benefits of physical activity interventions carry into wider life of people experiencing homelessness | Development of life and interpersonal skills | **High** (Group running[42], Group outdoor adventure[34])<br>**Moderate** (Soccer- tournament participation[36], Earn-a-bike[45], Group outdoor adventure[33])<br>**Low** (Soccer- group training and tournament participation[40], soccer- group training[35], Gardening group[43])<br>**Very low** (Group instructor-led dance[30]) |
|  | Improved social connection and building relationships with others | **High** (Group running[42], Group outdoor adventure[34])<br>**Moderate** (Soccer- tournament participation[36], Earn-a-bike[45], Group outdoor adventure[33])<br>**Low** (Soccer- group training and tournament participation[40], soccer- group training[35], Gardening group[43]) |
|  | Physical activity as a catalyst for positive healthy life change | **High** (Group outdoor adventure[34])<br>**Moderate** (Soccer- tournament participation[36])<br>**Low** (Soccer- group training and tournament participation[40], Gardening group[43]) |
|  | Practical and functional benefits developed from participation | **Moderate** (Earn-a-bike[45])<br>**Low** (Soccer- group training and tournament participation[40], soccer- group training[35]) |
| Perception of challenges related to physical activity participation whilst homeless | Homelessness presents specific barriers to PA participation | **High** (Group running[42])<br>**Moderate** (Earn-a-bike[45])<br>**Low** (Soccer- group training and tournament participation[40], soccer- group training[35])<br>**Very low** (Group instructor-led dance[30]) |
|  | Participating in soccer tournaments can be stressful | **High** (Soccer- group training and tournament participation[32])<br>**Moderate** (Soccer- tournament participation[36]) |
|  | Perceived poor performance/ aptitude can negatively impact confidence and coping | **High** (Soccer- group training and tournament participation[32])<br>**Moderate** (Soccer- tournament participation[36])<br>**Very low** (Group instructor-led dance[30]) |
| Belief of physical activity positive impact on self-medication, prescribed medication and addiction for people experiencing homelessness | Reduction in need for prescription medication and self-medication | **Moderate** (Earn-a-bike[45]) |
|  | Reduced substance misuse | **High** (Soccer- group training and tournament participation[32])<br>**Moderate** (Earn-a-bike[45])<br>**Low** (soccer- group training[35]) |
|  | Diversion from temptation of addiction | **Low** (soccer- group training[35]) |

| Quantitative findings |  |  |
| --- | --- | --- |
| Description of outcome and measurement tool | Outcome data | Level of confidence in evidence for intervention type |
| Positive bone health, measured from a blood sample and x-ray absorptiometry (DXA) in context of physical activity participation in people experiencing homelessness | Osteoclastin increased by 27% from pre to post intervention. This change was significantly different between the control and intervention groups ($p = 0.042$) | High (Soccer- group training[37]) |
| | Pre-post trunk bone mineral density increased by 1% ($p = 0.02$ in the intervention group. There was no difference between intervention and control groups. | High (Soccer- group training[37]) |
| No change to bone health, measured by x-ray absorptiometry (DXA), from participation in physical activity amongst people experiencing homelessness | Pre-post weight-bearing z-score increased from $0.6 \pm 1.1$ to $0.7 \pm 1.1$ ($p = 0.07$) in the intervention group. There was no difference between intervention and control groups. | High (Soccer- group training[37]) |
| | No pre-post changes in TRACP5b (bone resorption), plasma leptin or bone mineral density in intervention group ($p > 0.05$) | High (Soccer- group training[37]) |
| Positive changes to blood markers after participation in physical activity amongst people experiencing homelessness | LDL-cholesterol was lowered by $0.4 \text{ mmol L}^{-1}$ (95% CI: $-0.7$ to $-0.2$; $3.2 \pm 1.1$ to $2.8 \pm 0.8 \text{ mmol L}^{-1}$) in intervention group after 12 weeks, this change was significantly different to the control group ($p = 0.05$) | High (Soccer- group training[38]) |
| | HDL:LDL ratio increased by 0.06 (CI: 0.02 to 0.11) after 12 weeks in the intervention group ($0.43 \pm 0.13$ to $0.48 \pm 0.19$), which was different to the control group ($p = 0.05$) | High (Soccer- group training[38]) |

**Table 5:**
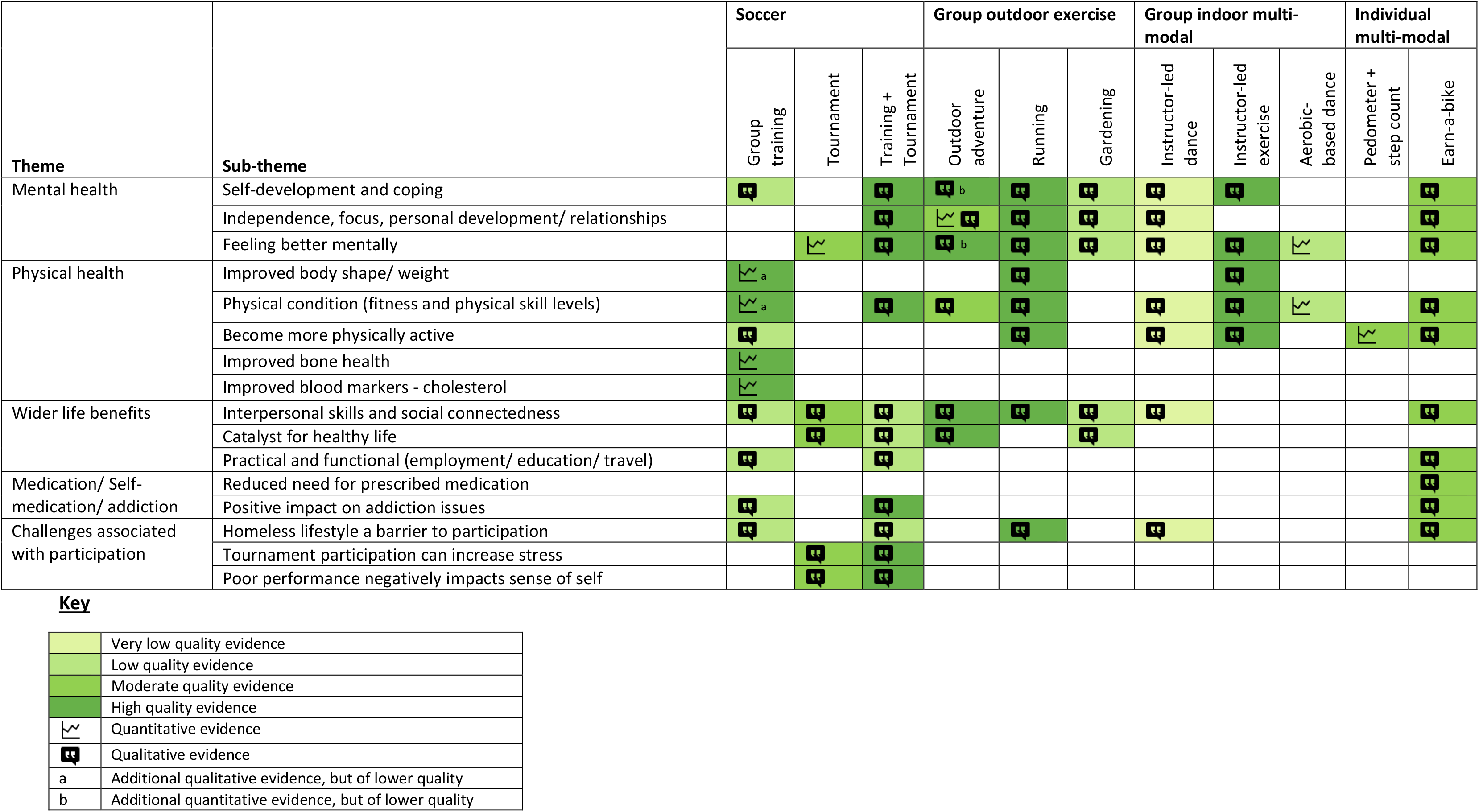
Summary of available evidence for physical activity interventions categorised by intervention type, findings and evidence quality.

### The impact of physical activity interventions on the mental health of PEH

There were several domains within mental health where both quantitative and qualitative evidence was synthesised, suggesting physical activity was beneficial (summarised in table 2). These included: enhanced confidence, empowerment, and self-esteem; resilience, coping and hope; independence, self-regulation and personal development; stress and anxiety; and mood and state of mind.

#### Enhanced confidence, empowerment, and self-esteem

There was high quality qualitative evidence that group running, soccer and indoor group exercise, and moderate quality qualitative evidence that group outdoor adventure and earn-a-bike enhanced confidence, empowerment, and self-esteem. However, the only quantitative study to assess outcomes in this domain used the Hope scale (agency sub-scale), finding no significant differences between groups. One soccer player suggested:

> “… Football gave me confidence and took away feelings of depression as it made me more social.”[32]

#### Resilience, coping and hope

There was high quality qualitative evidence that group running, and group outdoor adventure enhanced resilience, coping and hope. However, the only quantitative study to measure relevant outcomes using the Hope Scale (pathway domain) found no significant difference between intervention and control groups. A member of staff involved in delivering group outdoor adventure described changes in a participant’s ability to cope:

> “… when we went to Coniston, not even 10 min, we was there she wanted to come home, but she didn’t and she learned how to cope… she really enjoyed herself.”[47]

#### Independence, self-regulation and personal development

Qualitative evidence suggested that group running, and soccer (both high quality) and group outdoor adventure and earn-a-bike (both moderate quality) enhanced independence, self-regulation and personal development. This was supported by moderate quality quantitative evidence that outdoor adventure improved life functioning. An outdoor adventure participant describes how it impacted them:

> “when I leave here, I face any challenges… in my life, then I know that I will be able to do them because I’ve become a stronger person from coming here.”[33]

#### Stress and anxiety

There was high quality qualitative evidence that group running, indoor group exercise and outdoor adventure and moderate quality qualitative evidence that soccer and earn-a-bike had a positive effect on stress and anxiety. The studies that used quantitative measures to assess stress/ anxiety in soccer (moderate quality) and indoor group exercise (low quality) did not conclusively support the qualitative evidence. A participant at a gym-based programme said:

> “I… didn’t have the confidence to go outside, I felt a lot of like anxiety and this, the gym and stuff helps me with my anxiety really well.”[44]

#### Mood and state of mind

There was high quality qualitative evidence that soccer, group running and indoor group exercise and moderate quality qualitative evidence that earn-a-bike enhanced mood and state of mind. This was supported by moderate quality quantitative evidence that group outdoor adventure improved wellbeing.

### The impact of physical activity interventions on the physical health of PEH

Changes were shown in the following physical health domains: body shape and weight loss; fitness levels; physical skills development; and, physical activity levels. The synthesised findings are summarised in Table 3. Quantitative findings not corroborated by qualitative findings are summarised in Table 4.

#### Body shape and weight loss

Synthesised findings showed that indoor group exercise and group running (both high quality qualitative evidence) were perceived as improving body shape and facilitating weight loss, while soccer was shown to significantly decrease fat mass (high quality quantitative evidence).

> “I took my measurements when I started street fit, and I took my measurements now, and I’m a lot more buff.”[44]

#### Fitness levels

Synthesised findings for fitness levels showed that group running, group indoor training and earn-a-bike (all high-quality qualitative evidence) improved fitness levels, a finding backed up by a high quality quantitative study of soccer. A person who cycled with earn-a-bike described tryingto increase fitness:

> “… after riding, you know, for an hour, two hours, and sometimes I’ll ride for four hours. You know, I really want to make sure that my body is fit.”[45]

#### Physical skill development

While moderate quality qualitative evidence for group outdoor adventure was suggestive of positive changes physical skills development, the quantitative research exploring this domain through measuring postural balance showed no significant difference between intervention and control groups.

#### Physical activity levels

Synthesised findings showed that group indoor exercise and running groups (both high quality qualitative evidence) and earn-a-bike (moderate qualitative evidence) positively influenced physical activity levels. This was supported by a moderate quality quantitative pedometer and set a step count goal study. A woman from a running group described how since joining the group she now runs on her own:

> “I feel so much more body confident … I can actually run for the whole session without nearly dying. I also go out for runs on my own and I definitely think I’ve got faster.” [42]

#### Bone health and cholesterol

A high quality study measured markers of bone health[37] and cholesterol levels[38] in PEH who played soccer. Although not all bone markers improved, increases in osteoclastin from pre-to post-intervention were reported and this change was significantly different between controls and intervention groups. With regards to cholesterol markers [low-density lipoprotein-lipid (LDL)/ high-density lipoprotein (HDL)] cholesterol was lowered and LDL:HDL ratios increased in the intervention group after 12 weeks of soccer-findings which were significantly different (p=0.05) from the control group.

### Other considerations relevant to physical activity interventions for PEH

There were some findings relevant which described the impact of physical activity for PEH described in qualitative literature only. Themes include addiction, self-medication and medication; benefits carried into wider life; and, challenges to participation in physical activity when homeless (outlined in Table 4).

#### Addiction, self-medication and medication

Across several qualitative studies of soccer (high quality) and earn-a bike (moderate quality), physical activity positively influencing addiction was described. One person who played football stated:

> “I’m drinking less and do not think I need alcohol as much now… It’s great to feel this way and football is a focus for us.” [32]

#### Benefits of physical activity participation carried into wider life

Most of the qualitative studies, including soccer, running groups, earn-a-bike, outdoor adventure, gardening and dance, described benefits to wider life. Sub-themes included: development of life and interpersonal skills improved social connectedness and relationships with others, practical and functional benefits, and physical activity as a catalyst for positive healthy life change. A participant who undertook leisure centre-based group indoor training said:

> “I’ve noticed a massive improvement in my fitness, and it’s definitely keeping me motivated to live a healthy lifestyle, because you don’t put in all that hard work and then want to ruin it, you know what I mean?”[44]

… and similarly, how a participant of soccer described life change:

> “We can go back there and show that homelessness isn’t permanent and that you can change your life through sports.”[36]

#### Challenges to participation in physical activity when homeless

Qualitative evidence demonstrated the importance of acknowledging specific challenges related to physical activity PEH faced, which impacted uptake and dropout rates across a variety of interventions. Those who participated in soccer tournaments described heavy defeats impacting on self-worth.[32] Women who participated in running groups described lack of funds for transport or the unpredictability of homelessness as a barrier to attending.[48] There was also worry about loss of donated kit (e.g. running clothes)[48] and equipment (e.g. bicycle)[45] through theft and staff who led dance groups reported inconsistent attendance amongst shelter-dwellers.[30]

An overall summary of available evidence for physical activity interventions categorised by intervention type, findings and evidence quality is provided by Table 5.

## DISCUSSION

This review identified evidence for diverse physical activity interventions for PEH. The mixed methods methodology enabled a meaningfully configured synthesis of the breadth of available evidence. This review demonstrated positive impacts of physical activity for PEH in relation to mental and physical health outcomes with translation of benefits to wider life.

Physical activity interventions were heterogeneous, grouped into broad categories of soccer, group outdoor exercise, group indoor multi-modal exercise and individualised multimodal interventions. In terms of specific sports, soccer predominated (6/16). This is unsurprising considering its global resonance.[49] Two thirds (4/6) of soccer interventions included tournament participation. While benefits to tournament participation exist, negative experiences of pressure and fear of letting down teammates was qualitatively reported. Organisers of soccer tournaments for PEH should consider support to ameliorate impacts of possible pressure experienced, which could negatively impact mental health or self-management of addiction.

Group exercise appeared to be most beneficial for PEH. It is likely that group activities facilitated social support, especially pertinent for PEH whose social networks are often fragmented.[50] Configured qualitative and quantitative findings highlighted most evidence for mental health benefits in group outdoor exercise. Specifically, these benefits related to an improvement in mood and state of mind and increased independence, focus, personal development and ability to foster relationships. This may be related to emerging evidence for optimised benefits of outdoor exercise.[51] Corroboration of qualitative and quantitative evidence indicated that PEH who participated in physical activity interventions increased their physical activity levels. There is inherent difficulty comparing types of interventions for levels of benefit, as interventions and outcome measures were heterogeneous.

Many physical activity interventions included additional intervention components such as counselling, food, or sports kit. Consequently, it is not known if these additional components, enhanced or diluted the effect of physical activity. Moreover, descriptions of physical activity programme variables such as dosage were often lacking, limiting judgment of interventions.

Programme intensity deserves consideration. Soccer, which predominated, is a vigorous intensity sport [10 metabolic equivalent of task (METs) for competitive soccer and 7 METs for casual soccer][52] so it is likely this high entry level may be exclusionary for many PEH. Given the high prevalence and early manifestation of non-communicable diseases and poor general physical health in many PEH,[53] lower threshold physical activity interventions should be also considered. Some low threshold programmes were identified such as gardening and dance. People designing physical activity interventions for PEH should consider a range of abilities and likely poor physical condition, perhaps offering a choice of low threshold activity, as well as higher intensity options, depending on ability and interest.

Qualitative studies dominated the evidence base, justifying the methodological decision of a mixed methods review. The quality of evidence of most qualitative studies was judged to be high, with perspectives of staff enhancing credibility to the understanding of intervention impact. A limitation of the evidence identified was that only one quantitative study was an RCT. While, RCTs are considered the highest evidence level, this review attests to the usefulness of other study designs in this novel and emerging topic. It is acknowledged that RCTs may be especially difficult to undertake due to possible implementation barriers and complexities within this cohort. We propose that to build the evidence base, forms of controlled trials should be conducted where possible, with a view to including more randomised trials in the future. A further limitation was that feasibility outcomes such as adherence and retention rates were not described in many of the quantitative studies included in this review, though challenges to participation were described in several qualitative studies, therefore feasibility analysis and assessment of adherence and retention should be included in future studies.

Outcome measures employed were not consistent, e.g. cardiovascular fitness was measured in three different ways: the Yo-Yo endurance test, cycle ergometry and maximal treadmill testing. The evidence base is limited in terms of the most suitable outcome measures[54] to use in physical activity interventions for PEH. Future studies should explore the most suitable outcome measures with a view to improving consistency in their use to enable future evidence syntheses. [16:23].

Strengths of this review were its mixed methods design and the global spread of identified research including studies from the UK and Europe, North America and Australia. However, only high income countries were included, as low and middle income settings were considered to have different structural influences to homelessness, so a limitation of this systematic review is that the translation of findings to other settings is not known. The review team capitalised on expertise in inclusion health, physical activity interventions, evidence synthesis with input from expert medical librarians. Studies were quality assessed using a consistent ‘family’ of critical analysis tools from JBI.

## CONCLUSION

This mixed methods systematic review demonstrates the value in exploring literature across a wide variety of methodological domains to gain insights into the existence and impact of a variety of physical activity interventions for PEH. To confidently inform policy, more research in this topic is required, however, from a practice and research perspective, our results provide initial justification for the inclusion of this typically under-represented group in targeted physical activity interventions with benefits to multiple aspects of physical and mental health, and positive translation into wider life demonstrated. Future research should include larger-scale high quality quantitative research to provide more robust evidence regarding objective impact.

### OTHER INFORMATION

#### Protocol registration

This review was registered on PROSPERO, registration number: CRD42020216716. Found at https://www.crd.york.ac.uk/prospero/display_record.php?RecordID=216716. In the protocol we stated we would use Cochrane and Downs and Black risk of bias tools. However, once diversity of final studies were identified, the review team recognised that JBI risk of bias tools were more suited to studies within our review.

#### Support

JD was funded by a pre-doctoral fellowship from the National Institute for Health and Care Research (NIHR) School for Public Health Research (SPHR), Grant Reference Number PD-SPH-2015. The views expressed are those of the author(s) and not necessarily those of the NIHR or the Department of Health and Social Care.

#### Data availability

Search syntax, data extracted from included studies and data used for analysis are included in Supplementary files.

## Data Availability

All data produced in the present study are available upon reasonable request to the authors

## ACKNOWLEDGEMENTS

The authors would like to thank Jacqui Smith, clinical librarian at UCL for sharing her extensive knowledge and supporting the team with their protocol design, searching strategy and carrying out the searches. Thanks also to and David Mockler, librarian, Trinity College Dublin, Dr Cliona Ni Cheallaigh and Professor Andrew Hayward for their advice and support of this work.

