## Supplementary File 4 for "“You can change your life through sports”: Physical activity interventions to improve the health and wellbeing of adults experiencing homelessness: a mixed methods systematic review"

### Supplementary file 3: Physical activity interventions for people experiencing homelessness

| **Broad intervention theme** | **Physical activity intervention** | **Study** | **Number of participants receiving intervention and providing study data** | **Setting** | **Frequency** | **Intensity** | **Time** | **Type** | **Group/ individual** | **Presence of other components** |
| --- | --- | --- | --- | --- | --- | --- | --- | --- | --- | --- |
| Soccer | Soccer (group training) | Helge et al, 2014 &  Randers et al, 2012 | 18 | Soccer: Outdoors  four-a-side asphalt pitch (22× 16m)  Supervised training: fitness centre | Soccer: mean of 2.2 ± 0.7 sessions per week  Supervised training: mean of 0.5 ± 0.2 sessions per week, including 5 x strength exercises (one set of 10–12 repetitions) | Soccer: Verbally encouraged by coaches  Supervised training: low intensity warm up, moderate intensity strength training, exercises at > 15Repetition Maximum | Soccer: 10min warm up, duration of soccer training unclear  Supervised training: 15mins warm up + strength training time unclear | Soccer: 4 vs 4 Soccer games  Supervised training: cardiovascular warm up, strength-based resistance training | Group | NS |
|  |  | Sherry & Strybosch, 2012 | 144 | NS | Weekly | NS | 14min games with 1min half time break | Soccer | Group | Links to service |
|  | Soccer (Tournament participation) | Randers et al, 2018 | 15 | Outdoors  four-a-side artificial turf street soccer pitch (22× 16m with 4m wide goals) | NS | Majority of play at 70-100 %HR_peak_  Rate Perceived Exertion (x/10) 4.8± 2.5 | 4 day tournament.  Mean playing time per game: 11.1 ± 2.6min | Soccer | Group | International travel |
|  |  | Welty Peachey et al, 2013 | 11 | NS | NS | NS | 4 day tournament | Soccer | Group | Opening/ closing ceremonies, awards |
|  | Soccer (group training and Tournament participation) | Magee & Jeanes, 2013 | 6 | NS | NS | NS | 10 weeks training + tournament | Soccer | Group | Access to support services |
|  |  | Sherry, 2010 | 8 | NS | NS | NS | NS | Soccer | Group | Informal support, link to services |
| Group outdoor exercise | Group Outdoor Adventure | Norton et al, 2020 | 32 | Outdoors | NS | NS | NS | Archery, geocaching, rock climbing, and hiking | Group | Debrief, reflection on experience, individual and therapy therapy, childcare, ABC-R therapy model during outdoor activity |
|  |  | Parry 2021a | 30 | Countryside, 170km from usual residence | NS | NS | 4 days | Team based activities: canoeing, hiking, high ropes course, and raft building | Group | Preceding phase of 10 x Positive Youth Development life skills workshops |
|  |  | Parry 2021b | 54 | Outdoors | NS | NS | 3 to 4 days | canoeing, raft building, a mountain hike, high and low ropes courses, mountain biking, and caving. | Group | Preceding phase of 10 x Positive Youth Development life skills workshops and Team-based, structured reflections |
|  | Running groups | Dawes et al, 2019 | 11 | Local park | NS | Participant self-selected | 1 hour/ session | Running | Group | Running kit, healthy snack |
|  | Gardening groups | Grabbe et al, 2013 | 8 | Vegetable garden in rear parking lot of daytime shelter for  women | 2 x per week | NS | “staffed sessions” 2 hours, but women could  engage in gardening at any time during daylight  hours when the shelter was open. | Gardening, including: planting seeds,  pulling weeds, watering, harvesting, washing produce, | Group, but possible to undertake as an individual, if desired | Food preparation, health,  nutrition, and horticulture education |
| Group indoor multimodal exercise | Group instructor-led dance | Knestaut et al, 2010 | 11 | Recreation room of homeless shelter | Weekly | Gentle warm up, exercise to elevate heart rate and raise core temperature | 50 mins | Instructor-led group dance programme: stretching, cardiovascular exercise and dance genres including: hip hop, country-line dance, ballet, and creative movement/ improvisation | Group | Informal debrief |
|  | Group instructor-led, exercise | Malden et al, 2019 | 10 | Local leisure centre | Weekly | NS | Approx. 1 hour | instructor-led group fitness class: aerobic circuits and  strength-based resistance training exercises to music | Group | peer support  workshop and lunch |
|  | Sitting/ walking meditation, aerobic-based dance exercise training | Shors et al, 2014 | 8 | NS | 2x per week for 8 weeks | NS | 1 hour total: 20 mins Sitting meditation, 10 mins walking, 30mins  aerobic-based dance exercise training | Sitting meditation, walking, aerobic-based dance exercise training | Group | NS |
| Individual multimodal intervention | Pedometer with step count goals | Kendzor et al, 2017 | 15 | Shelter for homeless adults | NS | NS | 28 day period | Wear pedometer during waking ours and work towards 10,000 daily step goal | Individual | Educational newsletter, fruit/ veg snacks, |
|  | Earn-a-bike programme | Grimes & Smirnova, 2020 | 16 | Partnership between pedestrian and bicycling advocacy organization and local homeless outreach agency | NS | NS | NS | cycle safety training, including training on riding in heavy traffic and bicycle handling skills. Provision of bicycle and safety equipment | Unclear if training delivered in groups. Earning bicycle individual | Rules of the road and bicycle maintenance training |
