## Supplementary File 5 for "“You can change your life through sports”: Physical activity interventions to improve the health and wellbeing of adults experiencing homelessness: a mixed methods systematic review"

### Supplementary File 4: Study populations within studies identified in systematic review

| **Study** | **Description of participants** | **Number of Participants** | **Characteristics of participants who were homeless, does not include any participants who were staff** | | | | | | | |
| --- | --- | --- | --- | --- | --- | --- | --- | --- | --- | --- |
|  |  |  | **Age (years)** | **Educational achievement** | **Number in employment** | **Ethnicity/ race** | **Physical/Mental health/ psychological trauma** | **Biological Sex** | | |
|  |  |  |  |  |  |  |  | **F** | **M** | **Other** |
| Dawes et al, 2019 | Adult women with lived experience of homelessness, who had attended running groups for a minimum of 4 weeks. | 11 | Range: 23-57  Mean (SD): NS | NS | NS | NS | NS | 11 | 0 |  |
| Grabbe et al, 2013 | Adult women living in shelters or on the street. | 8 | Range: 20-59  Mean (SD): NS | 4/8 high school or less  4/8 some college or technical school | NS | NS | 4/8 ≥ 1 mental illnesses; including bipolar disorder, depression, obsessive-compulsive disorder, and borderline personality disorder | 8 | 0 |  |
| Grimes & Smirnova, 2020 | Adult men who had completed the "earn-a-bike" programme and had experienced homelessness at the time of participating in the programme. | 16 | Range: 30-65  Mean: 46.0  SD: NS | 1/16 less than high school  9/16 high school  4/16 some college  2/16 completed college | 2/8^b^ | 4/8 African American  4/8 White | NS | 0 | 16 |  |
| Helge et al, 2014 &  Randers et al, 2012 | Homeless adult men recruited from shelters and unemployment ofﬁces in Copenhagen. | 28 | Range: NS  Football group: Mean (SD):  36.4 (10.0)  Control group: Mean (SD):  42.7 (8.5) | NS | NS | NS | NS | 0 | 28 |  |
| Kendzor et al, 2017 | Adults living in transitional shelters, a score of ≥4 on Rapid Estimate of Adult Literacy in Medicine- Short Form, physically ambulatory and resident at shelter for ≤3 months. | 32 | Range: NS  Mean (SD): 48.38 (8.12) | 12.63 (2.08)^a^ | NS | 27/32 African American | NS | 8 | 24 |  |
| Knestaut et al, 2010 | Adult men and women living in a homeless shelter | 11 | Range: 18-50 | NS | NS | Caucasian (*n* = 9), Latina (*n* = 1) African American (*n* = 1) | NS | 8 | 3 |  |
| Magee & Jeanes, 2013 | Men who played in United Kingdom Homeless Word Cup squad. All of whom lived in either social services accommodation or hostels and had spent time living rough. | 6 | Range: 16-29  Mean (SD): NS | NS | 0/6 | NS | NS | 0 | 6 |  |
| Malden et al, 2019 | Adults experiencing homelessness (living in hostel accommodation or on the streets) and had attended ≤10 Street Fit Scotland sessions. | 10 | Range: 21-36  Mean (SD): NS | NS | 1/10 | NS | NS | 5 | 5 |  |
| Norton et al, 2020 | Adult women who, at the time of study, were living in a shelter for women without housing. | 82 | Range: 18-63  Mean (SD):  35.8 (11.4) | NS | NS | 41/83 Black  16/83 White  18/83 Hispanic  3/83 Other | NS | 82 | 0 |  |
| Parry et al, 2021a | Young people experiencing or at risk of homelessness who attended at least one My Strength Training for life (MST4Life) and consented/ engaged with qualitative data collection; housing service staff with one-to-one experience with MST4Life participants; and, Outdoor Adventure Education (OAE) staff supporting phase 2 of MST4Life programme. | 30 PEH  6 housing service staff  5 OAE staff | Range: 16-24  Mean (SD):  20 (2.04) | NS | 10/30 | 18/30^c^ White  8/30^c^ Mixed ethnicities  2/30^c^ Black  2/30 ^c^ Asian  1/30^c^ other | 23/30^c^ mental health condition | 15 | 14 | 1^d^ |
| Parry et al, 2021b | Young people experiencing or at risk of homelessness who had participated in phase 1 of My Strength Training for life (MST4Life). | 54 | Range: 16-24  Mean, SD:  19.43 (2.18) | NS | 3/54*** | 33/54^c^ White  11/54^c^ multiple or mixed ethnic groups  2/54^c^ Asian  2/54^c^ Black  2/54^c^ Other | NS | 22 | 31 | 1^d^ |
| Randers et al, 2018 | Adult women participating in the Women’s Homeless World Cup. | 15 | Range: NS  Mean, SD: 30.3 ± 5.0 | NS | NS | NS | NS | 15 | 0 |  |
| Shors et al, 2014 | Adult women (mothers) who were recently homeless and >1 month poverty, trauma and addictive behaviour | 15 | Range: 18-36  Mean (SD):  25.0 (5.0) | NS | NS | NS | NS | 15 | 0 |  |
| Sherry, 2010 | Men, >16 years old, who played in Australian 2006 Homeless World Cup team, had experienced homelessness in previous 2 years and/or in a drug or alcohol rehabilitation programme. | 8 | NS | NS | NS | NS | NS | 0 | 8 |  |
| Sherry & Strybosch, 2012 | Soccer program participants of the Community Street Soccer Programme; coaches and support workers of the programme. | 165 players  11 Coaches  10 (support workers) | Range: ‘under 18’-40  Most concentrated cohort 20-24  Mean (SD): NS | NS | NS | NS | 66/165^c^ suffering from depression or other mental illness | 23^c^ | 142^c^ |  |
| Welty Peachey et al, 2013 | Participants were players, coaches and administrators  from Street Soccer USA Cup (SSUSA) who represented the geographic areas served by SSUSA | 11 players  6 staff | Range: 17-54  Mean (SD): NS | NS | NS | 4/11 White  4/11 African-American  2/11 Hispanic  1/11 Mixed race | NS | 3 | 8 |  |

NS: not stated, SD: standard deviation, ^a^ mean(sd) years of education, ^b^ number in employment/education/training, ^c^ percentage supplied without accompanying numbers, closest whole number deduced relative to number of participants is reported, ^d^ identified as transgender
