## Supplementary File 1 for "“You can change your life through sports”: Physical activity interventions to improve the health and wellbeing of adults experiencing homelessness: a mixed methods systematic review"

### Supplementary file 1: Search syntax

This document contains the search syntax used in Medline, EMBASE, Web of Science, CINAHL, SPORTDiscus, the Cochrane Library and PsycINFO

#### Database: Ovid MEDLINE(R) and Epub Ahead of Print, In-Process & Other Non-Indexed Citations and Daily <1946 to December 14, 2020>

Search Strategy:

--------------------------------------------------------------------------------

1	homeless persons/ or homeless youth/

2	(homeless person* or homeless* or street dwell* or rough sleep* or no fixed abode or temporary accommodation or temporary housing or insecure housing or hostel).tw.

3	exp Exercise/

4	(physical activit* or (bik* or bicycl* or cardio* cycl* or danc* fit* or garden* or jog* or run* or swim* or walk* or zumba or salsa or pilates or yoga or tai chi)).tw.

5	exp Sports/

6	(sport* or (badminton or boxing or ball or gym* or soccer or tennis)).tw.

7	(exercis* or ((exercise* adj therapy) or rehab*)).tw.

8	((physical adj activ*) or educa* or train*).tw.

9	((aerobic* or cadio* or physi*) adj (exercise* or rehab* or therap* or fit* or train*)).tw

10	((resistance or strength* or weight*) adj (train* or exercis*)).tw.

11	Resistance Training/

12	(keep* adj (active or fit)).tw.

13	(High Intensity Interval Training or HIIT or (circuit* adj train*)).tw.

14	High-Intensity Interval Training/

15	Circuit-Based Exercise/

16	Pliability/

17	Anxiety/

18	Depression/

19	Accidental Falls/

20	Frailty/

21	"Quality of Life"/

22	(Balance or flexibility or strength or anxiety or depression or (exercise adj6 (tolerance or capacity)) or falls or (physical adj (fitness or function)) or frailty or function or independence or mood or quality of life or wellness or wellbeing).tw.

23	1 or 2

24	3 or 4 or 5 or 6 or 7 or 8 or 9 or 10 or 11 or 12 or 13 or 14 or 15

25	16 or 17 or 18 or 19 or 20 or 21 or 22

26	23 and 24 and 25

#### Database: Embase <1980 to 2021 Week 05>

Search Strategy:

--------------------------------------------------------------------------------

1	homeless person/ or homeless youth/ (2250)

2	(homeless person* or homeless* or street dwell* or rough sleep* or no fixed abode or temporary accommodation or temporary housing or insecure housing or hostel).tw. (14412)

3	exp exercise/ (347510)

4	(physical activit* or (bik* or bicycl* or cardio* cycl* or danc* fit* or garden* or jog* or run* or swim* or walk* or zumba or salsa or pilates or yoga or tai chi)).tw. (666192)

5	exp sport/ (167693)

6	(sport* or (badminton or boxing or ball or gym* or soccer or tennis)).tw. (143093)

7	(exercis* or ((exercise* adj therapy) or rehab*)).tw. (603637)

8	((physical adj activ*) or educa* or train*).tw. (1520464)

9	((aerobic* or cadio* or physi*) adj (exercise* or rehab* or therap* or fit* or train*)).tw. (99728)

10	((resistance or strength* or weight*) adj (train* or exercis*)).tw. (25991)

11	resistance training/ (19850)

12	(keep* adj (active or fit)).tw. (298)

13	(High Intensity Interval Training or HIIT or (circuit* adj train*)).tw. (2875)

14	high intensity interval training/ (2645)

15	circuit training/ (250)

16	pliability/ (3082)

17	anxiety/ (211042)

18	depression/ (371712)

19	falling/ (42141)

20	frailty/ (13530)

21	"quality of life"/ (494839)

22	(Balance or flexibility or strength or anxiety or depression or (exercise adj6 (tolerance or capacity)) or falls or (physical adj (fitness or function)) or frailty or function or independence or mood or quality of life or wellness or wellbeing).tw. (4244181)

23	1 or 2 (14871)

24	3 or 4 or 5 or 6 or 7 or 8 or 9 or 10 or 11 or 12 or 13 or 14 or 15 (2548913)

25	16 or 17 or 18 or 19 or 20 or 21 or 22 (4530822)

26	23 and 24 and 25 (634)

#### Databases: Web of Science, CINAHL, Cochrane and SportsDISCUS Search strategy

"homeless person*" or homeless* or "street dwell*" or "rough sleep*" or "no fixed abode" or "temporary accommodation" or "temporary housing" or "insecure housing" or hostel

And

“physical activit*” or bik* or bicycl* or cardio* cycl* or “danc* fit*” or garden* or jog* or run* or swim* or walk* or zumba or salsa or pilates or yoga or “tai chi”or sport* or badminton or boxing or ball or gym* or soccer or tennis or exercis* or “exercise* adj1 therapy” or rehab*or “physical adj1 activ*” or educa* or train* or aerobic* or cadio* or “physi* adj1 exercise*” or rehab* or therap* or fit* or train* or resistance or strength* or “weight* adj1 train*” or exercis* or “keep* adj1 active” or fit or “High Intensity Interval Training” or HIIT or “circuit* adj1 train*”

And

Balance or flexibility or strength or anxiety or depression or “exercise adj6 tolerance” or capacity or falls or “physical adj fitness” or “physical adj function” or frailty or function or independence or mood or “quality of life” or wellness or wellbeing

#### Database: APA PsycInfo <1806 to February Week 2 2021>

Search Strategy:

--------------------------------------------------------------------------------

1	("homeless person*" or homeless* or "street dwell*" or "rough sleep*" or "no fixed abode" or "temporary accommodation" or "temporary housing" or "insecure housing" or hostel).tw

2	exp Exercise/

3	exp Sports/

4	(physical activit* or (bik* or bicycl* or cardio* cycl* or danc* fit* or garden* or jog* or run* or swim* or walk* or zumba or salsa or pilates or yoga or tai chi)).tw.

5	(sport* or (badminton or boxing or ball or gym* or soccer or tennis)).tw.

6	(exercis* or ((exercise* adj therapy) or rehab*)).tw.

7	((physical adj activ*) or educa* or train*).tw.

8	((aerobic* or cadio* or physi*) adj (exercise* or rehab* or therap* or fit* or train*)).tw.

9	((resistance or strength* or weight*) adj (train* or exercis*)).tw.

10	(keep* adj (active or fit)).tw.

11	(High Intensity Interval Training or HIIT or (circuit* adj train*)).tw.

12	Anxiety/

13	"Depression (Emotion)"/

14	Falls/

15	"Quality of Life"/

16	(Balance or flexibility or strength or anxiety or depression or (exercise adj6 (tolerance or capacity)) or falls or (physical adj (fitness or function)) or frailty or function or independence or mood or quality of life or wellness or wellbeing).tw.

17	2 or 3 or 4 or 5 or 6 or 7 or 8 or 9 or 10 or 11

18	12 or 13 or 14 or 15 or 16

19	1 and 17 and 18
