## Supplementary File 2 for "“You can change your life through sports”: Physical activity interventions to improve the health and wellbeing of adults experiencing homelessness: a mixed methods systematic review"

Supplementary Table 2a: Quality assessment of qualitative studies, using JBI critical appraisal checklists

|  | Qualitative | | | | | | | | | | |
| --- | --- | --- | --- | --- | --- | --- | --- | --- | --- | --- | --- |
|  | Dawes 2019 | Grabbe 2013 | Grimes 2020 | Knestaut 2010 | Malden 2019 | Magee 2013 | Parry 2021a | Parry 2021b | Sherry 2010 | Sherry 2012 | Welty 2013 |
| Q1: Is there congruity between the stated philosophical perspective and the research methodology? | Yes | Yes | Yes | No | Yes | Yes | Yes | Yes | Unclear | Yes | Yes |
| Q2: Is there congruity between the research methodology and the research question or objectives? | Yes | Yes | Yes | No | Yes | Yes | Yes | Yes | Unclear | Yes | Yes |
| Q3: Is there congruity between the research methodology and the methods used to collect data? | Yes | Yes | Yes | No | Yes | Yes | Yes | Yes | Yes | Yes | Yes |
| Q4: Is there congruity between the research methodology and the representation and analysis of data? | Yes | No | Yes | No | Yes | Yes | Yes | Yes | Yes | Yes | Yes |
| Q5: Is there congruity between the research methodology and the interpretation of results? | Yes | No | Yes | Unclear | Yes | Yes | Yes | Yes | Yes | Yes | Yes |
| Q6: Is there a statement locating the researcher culturally or theoretically? | Yes | No | No | Yes | Yes | Yes | No | Yes | No | No | No |
| Q7: Is the influence of the researcher on the research, and vice- versa, addressed? | No | No | No | No | Yes | Yes | No | Yes | No | No | Unclear |
| Q8: Are participants, and their voices, adequately represented? | Yes | Yes | Yes | No | Yes | Yes | Yes | Yes | Yes | Yes | Yes |
| Q9: Is the research ethical according to current criteria or, for recent studies, and is there evidence of ethical approval by an appropriate body? | Yes | Yes | Yes | No | Yes | Yes | Yes | Unclear | Unclear | No | No |
| Q10: Do the conclusions drawn in the research report flow from the analysis, or interpretation, of the data? | Yes | Yes | Yes | Unclear | Yes | Yes | Yes | Yes | Yes | Yes | Yes |
| Dependability rating (Based on number of “yes” across Q2, 3, 4, 6 and 7)[1] | 4/5  Unchanged | 2/5  Downgrade one level | 3/5 Downgrade one level | 1/5  Downgrade two levels | 5/5  Unchanged | 5/5 Unchanged | 3/5 Downgrade one level | 5/5 Unchanged | 2/5 Downgrade one level | 3/5 Downgrade one level | 3/5 Downgrade one level |
| Credibility rating (Based on review of findings)[1] | Unequivocal | Equivocal | Unequivocal | Equivocal | Unequivocal | Unequivocal | Unequivocal | Unequivocal | Equivocal | Equivocal | Unequivocal |
| High, mod, low, v low | High | Low | Moderate | Very Low | High | High | Moderate | High | Low | Low | Moderate |

1.	Munn, Z., et al., *Establishing confidence in the output of qualitative research synthesis: the ConQual approach.* BMC Medical Research Methodology, 2014. **14**(1): p. 108.

Supplementary Table 2b: Quality assessment of quantitative studies, using JBI critical appraisal checklists

|  | RCT | Quasi-experimental | | | | Analytical Cross-sectional |
| --- | --- | --- | --- | --- | --- | --- |
|  | Kendzor 2017 | Shors 2014 | Helge 2014 | Randers 2012 | Norton 2020 | Randers 2018 |
| Was true randomization used for assignment of participants to treatment groups? | Yes |  |  |  |  |  |
| Was allocation to treatment groups concealed? | Unclear |  |  |  |  |  |
| Were treatment groups similar at the baseline? | Yes |  |  |  |  |  |
| Were participants blind to treatment assignment? | No |  |  |  |  |  |
| Were those delivering treatment blind to treatment assignment? | No |  |  |  |  |  |
| Were outcomes assessors blind to treatment assignment? | Unclear |  |  |  |  |  |
| Were treatment groups treated identically other than the intervention of interest? | No |  |  |  |  |  |
| Was follow up complete and if not, were differences between groups in terms of their follow up adequately described and analyzed? | Yes | Unclear | No | No | No |  |
| Were participants analyzed in the groups to which they were randomized? | Yes |  |  |  |  |  |
| Were outcomes measured in the same way for treatment groups? | Yes |  |  |  |  |  |
| Were outcomes measured in a reliable way? | Yes | Unclear | Yes | Yes | Yes | Yes |
| Was appropriate statistical analysis used? | Yes | Unclear | Yes | Yes | Yes | Yes |
| Was the trial design appropriate, and any deviations from the standard RCT design accounted for in the conduct and analysis of the trial? | Yes |  |  |  |  |  |
| Is it clear in the study what is the ‘cause’ and what is the ‘effect’ (i.e. there is no confusion about which variable comes first)? |  | Yes | Yes | Yes | Yes |  |
| Were the participants included in any comparisons similar? |  | Unclear | Yes | Yes | Yes |  |
| Were the participants included in any comparisons receiving similar treatment/care, other than the exposure or intervention of interest? |  | Yes | Yes | Yes | No |  |
| Was there a control group? |  | Yes | Yes | Yes | Yes |  |
| Were there multiple measurements of the outcome both pre and post the intervention/exposure? |  | No | No | No | No |  |
| Were the outcomes of participants included in any comparisons measured in the same way? |  | Unclear | Yes | Yes | Yes |  |
| Were the criteria for inclusion in the sample clearly defined? |  |  |  |  |  | Unclear |
| Were the study subjects and the setting described in detail? |  |  |  |  |  | Yes |
| Was the exposure measured in a valid and reliable way? |  |  |  |  |  | Yes |
| Were objective, standard criteria used for measurement of the condition? |  |  |  |  |  | Yes |
| Were confounding factors identified? |  |  |  |  |  | No |
| Were strategies to deal with confounding factors stated? |  |  |  |  |  | No |
| Quantitative quality rating: [2]  (High: >75%/ Mod: 50-75%/ Low: 25-50%/ v low: <25%) | 8/13 = 61.5%  Mod | 3/9 = 33.3%  Low | 7/9 = 77.7%  High | 7/9 = 77.7%  High | 6/9 = 66.7%  Mod | 5/8 = 62.5%  Mod |

2. Munn, Zachary; Barker, Timothy Hugh; Moola, Sandeep; Tufanaru, Catalin; Stern, Cindy; McArthur, Alexa; Stephenson, Matthew; Aromataris, Edoardo. Methodological quality of case series studies: an introduction to the JBI critical appraisal tool. JBI Evidence Synthesis 18(10):p 2127-2133, October 2020. | DOI: 10.11124/JBISRIR-D-19-00099
