## Supplementary File 3 for "“You can change your life through sports”: Physical activity interventions to improve the health and wellbeing of adults experiencing homelessness: a mixed methods systematic review"

| **Overarching theme** | **Theme** | **Sub theme** | **Level of confidence in evidence for intervention type** |
| --- | --- | --- | --- |
| Physical health benefits of physical activity interventions for PEH | Belief that physical activity improves the body shape of people experiencing homelessness | Improved body shape and self-image | **High** (Running groups[1], Indoor group instructor-led exercise[2]) |
|  |  | Weight loss | **High** (Running groups[1])  **Low** (Soccer- group training[3]) |
|  | Belief that physical activity improves physical condition of people experiencing homelessness | Perception of improved fitness levels | **High** (Running groups[1], Soccer- group training and Tournament participation[4], Indoor group instructor-led exercise[2])  **Moderate** (Earn-a-bike[5])  **Low** (Soccer- group training[3])  **Very Low** (Group instructor-led dance[6]) |
|  |  | Physical skill development | **Moderate** (Group Outdoor adventure[7])  **Low** (Soccer- group training[3])  **Very Low** (Group instructor-led dance[6]) |
|  | Belief that participating in physical activity interventions makes people experiencing homelessness more active in general | Participating in the physical activity intervention increased physical activity in everyday life | **High** (Running groups[1], Indoor group instructor-led exercise[2])  **Moderate** (Earn-a-bike[5])  **Low** (Soccer-group training[3]) |
|  |  | Broader aspects of the intervention (clothes, equipment, skill) facilitated physical activity participation | **High** (Running groups[1])  **Very Low** (Group instructor-led dance[6]) |
| Mental health benefits of physical activity interventions for PEH | Belief that physical activity facilitated people experiencing homelessness in self-develop and their ability to cope with life situations | Confidence, empowerment and self-esteem | **High** (Group running[1], Soccer- group training and tournament participation[4], Indoor group instructor-led exercise[2])  **Moderate** (Earn-a-bike[5], Group outdoor adventure[7])  **Low** (Gardening group[8], Soccer- group training[3])  **Very low** (Group instructor-led dance[6]) |
|  |  | Resilience, coping and hope | **High** (Group running[1], Group outdoor adventure[9])  **Moderate** (Group outdoor adventure[7])  **Low** (Gardening group[8], Soccer- group training[3])  **Very low** (Group instructor-led dance[6]) |
|  | Belief that physical activity resulted in people experiencing homelessness feeling mentally better | Independence, focus, personal development and relationships | **High** (Group running[1], Soccer- group training and tournament participation[4])  **Moderate** (Group outdoor adventure[7], Earn-a-bike[5])  **Low** (Gardening group[8], Soccer- group training and tournament participation [10])  **Very low** (Group instructor-led dance[6]) |
|  |  | Positive effect on stress and anxiety | **High** (Group running[1], Indoor group instructor-led exercise[2], Group outdoor adventure[9])  **Moderate** (Soccer- tournament participation[11], Earn-a-bike[5])  **Low** (Gardening group[8], Soccer- group training and tournament participation [10])  **Very low** (Group instructor-led dance[6]) |
| The impact of physical activity interventions on the wider life of PEH | Belief that the benefits of physical activity interventions carry into wider life of people experiencing homelessness | Development of life and interpersonal skills | **High** (Group running[1], Group outdoor adventure[9])  **Moderate** (Soccer- tournament participation[11], Earn-a-bike[5], Group outdoor adventure[7])  **Low** (Soccer- group training and tournament participation[10], soccer- group training[3], Gardening group[8])  **Very low** (Group instructor-led dance[6]) |
|  |  | Improved social connection and building relationships with others | **High** (Group running[1], Group outdoor adventure[9])  **Moderate** (Soccer- tournament participation[11], Earn-a-bike[5], Group outdoor adventure[7])  **Low** (Soccer- group training and tournament participation[10], soccer- group training[3], Gardening group[8]) |
|  |  | Physical activity as a catalyst for positive healthy life change | **High** (Group outdoor adventure[9])  **Moderate** (Soccer- tournament participation[11])  **Low** (Soccer- group training and tournament participation[10], Gardening group[8]) |
|  |  | Practical and functional benefits developed from participation | **Moderate** (Earn-a-bike[5])  **Low** (Soccer- group training and tournament participation[10], soccer- group training[3]) |
|  | Perception of challenges related to physical activity participation whilst homeless | Homelessness presents specific barriers to PA participation | **High** (Group running[1])  **Moderate** (Earn-a-bike[5])  **Low** (Soccer- group training and tournament participation[10], soccer- group training[3])  **Very low** (Group instructor-led dance[6]) |
|  |  | Participating in soccer tournaments can be stressful | **High** (Soccer- group training and tournament participation[4])  **Moderate** (Soccer- tournament participation[11]) |
|  |  | Perceived poor performance/ aptitude can negatively impact confidence and coping | **High** (Soccer- group training and tournament participation[4])  **Moderate** (Soccer- tournament participation[11])  **Very low** (Group instructor-led dance[6]) |
|  | Belief of physical activity positive impact on self-medication, prescribed medication and addiction for people experiencing homelessness | Reduction in need for prescription medication and self-medication | **Moderate** (Earn-a-bike[4]) |
|  |  | Reduced substance misuse | **High** (Soccer- group training and tournament participation[10])  **Moderate** (Earn-a-bike[4])  **Low** (soccer- group training[7]) |
|  |  | Diversion from temptation of addiction | **Low** (soccer- group training[7]) |
